# Effective Coverage of Management of Wasting in Ethiopia

**DOI:** 10.1101/2024.04.23.24306206

**Authors:** Alinoor Mohammed Farah, Samson Gebremedhin, Beshada Rago, Aweke Kebede, Kemeria Barsenga, Mufaro Chiriga, Tefara Darge, Tafara Ndumiyana, Tayech Yimer, Hiwot Darsene, Shibru Kelbessa, Beza Yilma, Seifu Hagos Gebreyesus

**Affiliations:** School of Public Health, College of Health Sciences, Addis Ababa University, Addis Ababa, Ethiopia; World Food Programme (WFP), Addis Ababa, Ethiopia; Nutrition Coordination Office, Federal Ministry of Health, Ethiopia; Department of Public Health Nutrition, School of Public Health, College of Medicine and Health Sciences, Jigjiga University, Jigjiga, Ethiopia

**Keywords:** Ethiopia, Wasting, Contact coverage, Quality-adjusted coverage

## Abstract

**Background:** Child wasting, or acute malnutrition, is a life-threatening condition that increases the risk of death and serious illness. Despite efforts such as the Global Action Plan on Child Wasting, which aims to reduce wasting prevalence to less than 3% by 2030, challenges persist, with Ethiopia recording a 7.2% rate. A major shortcoming of the global strategy is the focus on contact coverage, which often overlooks service quality. Effective coverage that incorporates the quality of health services offers a solution.

**Objective:** To assess the effective coverage of management of child wasting in six regions of Ethiopia.

**Data and Methods:** We conducted a secondary analysis of cross-sectional data obtained from household and institutional surveys. Participants included caregivers and children aged 6-59 months. By combining household data with expanded measures of health facility readiness and process quality from health posts, we calculated the quality-adjusted coverage.

**Results:** Contact coverage for severe acute malnutrition (SAM) and moderate acute malnutrition (MAM) was 40% and 37%, respectively. Readiness scores for providing SAM and MAM services at health posts were 57.9% and 76.4%, respectively. The input-adjusted coverage for SAM and MAM, considering facility readiness, was 23% and 28%, respectively. The coverage adjusted for complete intervention receipt was 7% for SAM and 12% for MAM. Quality-adjusted coverage for both SAM and MAM was 4%.

**Conclusion:** Efforts to address acute malnutrition in the Ethiopian health system show commendable progress but also highlight critical gaps and inconsistencies. A holistic, quality-driven approach is needed to effectively combat child-wasting in Ethiopia.

**Strengths and limitations of this study:**

- Household surveys and facility data were concurrently collected within the same year. This allowed for an effective comparison between the readiness of facilities and services provided for acute malnutrition at that time.
- The selection of items for readiness and process quality was guided by WHO Service Availability and Readiness Assessment and the National Guideline for the Management of Acute Malnutrition.
- The study included only health posts in selected IMAM districts. This restricts the generalizability of the findings, as the care characteristics and quality at these excluded facilities might differ significantly.
- Although the health facility survey was extensive, it did not capture all the necessary data for a holistic calculation of the care cascade, particularly missing information needed for user adherence-adjusted coverage and outcome-adjusted coverage.

## Introduction

Childhood malnutrition remains a pressing public health concern because of its intrinsic association with morbidity and mortality. Globally, malnutrition, particularly in its severe forms, demands increased emphasis on effective and widespread interventions[1]. Current evidence has highlighted that with the systematic delivery of a core set of 19 maternal and child interventions, there is the potential to mitigate maternal and neonatal deaths and stillbirths by almost a quarter[2]. Furthermore, facilitating access to a mere ten evidence-based nutrition interventions can lead to a 15% reduction in mortality rates among children under five[3].

The Global Action Plan on Child Wasting underscores the imperative nature of enhancing the coverage and efficiency of interventions focused on both preventing and treating wasting. This commitment aligns with the ambitious Sustainable Development Goal (SDG) to reduce the prevalence of wasting to less than 3% by 2030[4]. Despite the progress, 7.2% of Ethiopian children remain wasted[5], underscoring the persistent challenge in the face of improvements.

However, for the actualization of these global ambitions, it is paramount to recognize the dual necessity of not only expansive reach but also robust quality of interventions. A pivotal limitation of the prevailing global approach is its primary focus on contact coverage indicators, often sidelining the crucial aspect of service quality[6]. Such an approach provides only a tentative link to real-world health benefits for the population in need[7]. Introducing the concept of effective coverage helps bridge this gap. Defined as the proportion of potential health gain that is effectively realized, effective coverage emphasizes ensuring that individuals receive timely and high-quality health services, guaranteeing the desired health outcomes[8].

Over the last two decades, Ethiopia has demonstrated significant commitment to this global ambition. Strenuous efforts have been channeled to guarantee that all Ethiopian children benefit from equal access to nutrition services, including prevention and management of acute malnutrition, facilitated not only through health facilities but also through community-based programs. Instrumental in this endeavor has been the National Health Extension Program (HEP), a landmark initiative that has remarkably augmented service coverage, especially in the remote and rural terrains of the nation[9].

Nevertheless, while the strides are commendable, the journey is not without challenges. Disparities in service coverage, coupled with inequities in healthcare accessibility, are significant obstacles to realizing the full potential of nutritional interventions aimed at child health and well-being[10]. Furthermore, the sporadic nature of household visits and poor quality of growth monitoring for children by health extension workers (HEWs) speaks to the inconsistency in the quality of services rendered[11].

Effective coverage is a method for assessing the degree to which a service is utilized while considering its quality. To determine effective coverage, a simple approach is to multiply the percentage of the population using the service by a measure of its quality. However, a more comprehensive method for calculating the effective coverage was recently proposed. The seven-step coverage framework starts with the intended client population and tracks hypothetical progression through a series of health benefit losses at various stages. This starts with an individual in need of contact with a health service, followed by input-adjusted coverage where the service is ready, intervention coverage where health services are received, quality-adjusted coverage where the services are provided according to standards, user adherence-adjusted coverage where user adherence is considered, and finally, outcome-adjusted coverage where the health gain achieved is considered[7, 12].

While the notion of effective coverage holds significance, most studies addressing this area have centered on antenatal care (ANC), postnatal care, facility delivery, family planning, HIV, sick childcare, and growth monitoring[13–20]. Notably, investigations focusing specifically on the management of wasting remain scarce. In the Ethiopian context, past research has conducted facility-based evaluations of the coverage and quality of nutrition-specific interventions [21]. This study emphasizes a substantial knowledge gap regarding effective coverage in the realm of wasting management. With this backdrop, our study aims to shed light on the effective coverage management of wasting in Ethiopia, utilizing insights from household and health facility surveys, and bridging the existing research gap.

## Methods

### Study setting

Within Ethiopia’s primary healthcare framework, the system is predominantly structured around primary hospitals, health centers, and five elite health posts. Health posts are staffed by two health extension workers, collectively serving an estimated to 4,000-5,000 rural residents. Launched in 2003, the Ethiopian Health Extension Program (HEP) was instrumental in aiming for universal primary healthcare coverage of the country’s rural demographics. This systematic program, facilitated by health extension workers, delivers essential promotive, preventive, and curative services through a combination of outreach initiatives and health post-based methods.

The management of acute malnutrition, particularly Severe Acute Malnutrition (SAM) and Moderate Acute Malnutrition (MAM), is a critical responsibility of Health Extension Workers (HEWs) at the post-health level in Ethiopia. Historically, the focus has been on SAM management, but in 2019, a new guideline was introduced that integrated the management of MAM into the health system to improve continuity of care, which was previously managed by the Ethiopian National Disaster Reduction and Management Commission (ENDRMC). This guideline, known as the Integrated Management of Acute Malnutrition (IMAM), mandates that HEWs manage MAM, including the distribution of fortified foods to beneficiaries. Currently, IMAM is implemented in approximately 150 priority districts across the Afar, Amhara, Oromia, Somali, SNNP, and Sidama regions, as identified by the Federal Ministry of Health (FMOH). This study is part of a broader research initiative funded by the World Food Programme (WFP) aimed at evaluating the impact of IMAM on program coverage and effectiveness. The goal is to enhance the initiative by addressing implementation challenges and promoting the best practices.

The project encompasses 18 districts distributed across six regions in Ethiopia. It adopts a longitudinal study design that involves 12 districts that implement IMAM and six districts that do not (**Supplementary Table 1**). The first round of surveys was conducted in June 2023. The 12 IMAM implementing districts were divided into two groups, with each group being a block. One group will receive active support from Implementation Science (IS), while the other group will continue with the usual implementation of IMAM without any support from IS.

### Study design and data source

For this evaluation, a community-based survey was conducted in June 2023. This study presents a secondary analysis of baseline cross-sectional data, particularly from IMAM-implementing districts, encompassing both household and institutional surveys. The latter assessed the readiness of health posts and the provision of acute malnutrition services across 72 health posts in six regions (12 health posts per region).

### Participants

This survey included all caregivers and children aged 6–59 months who resided in 12 of the 18 study districts, particularly the IMAM-implementing districts. It also includes all health posts with one or two health extension workers per health post serving the families.

### Sample size

The sample size for the baseline survey was determined based on assumptions regarding the calculations, prevalence of malnutrition, and screening coverage for children under five. These assumptions included a power of 80%, baseline screening coverage, and malnutrition prevalence of 50% and 16%, respectively, a prevalence ratio of 1.22, indicative of a small effect size, a design effect of 1.5, and a 10% non-response rate. The total sample size for the study was 2,160 children aged 6-59 months. Of these, 1,440 children aged 6-59 months were included in the study, and health facility assessments were conducted in IMAM-implementing districts. In addition, 72 health posts were randomly selected, and one or two health extension workers from each health post serving the population of the study clusters were included.

### Sampling

Study participants were selected using a multistage cluster sampling approach. Initially, in each district, three kebeles (the smallest administrative units in Ethiopia) representing three different agro-ecologies were selected using stratified sampling. In kebeles with active 1-30 networks, we first listed all existing networks and randomly selected five networks; then, eight children were randomly included. In kebeles that did not have 1-30 networks, we exhaustively listed the villages/ “gots” in that kebele, randomly sampled two villages, and in each selected village, 20 children were selected using the Random Walk Method [22]. In both the methods, 40 children were drawn per kebele.

### Data collection

Data were collected using pretested tools prepared in local languages (Amharic, Afan Oromo, and Somali). Survey data were gathered using the Open Data Kit (ODK), an open-source and user-friendly application system on which SPH has the capability to leverage. This ODK platform ensured close-to-real-time quality data collection, cleaning, and monitoring. The data were uploaded daily by enumerators to the ODK cloud server.

Survey data were collected by 30 trained and experienced enumerators and six supervisors. Recruitment of personnel was made based on multiple criteria including educational status (at least diploma holders in health-related disciplines), experience in similar surveys, previous experience of collecting data using ODK, and successful completion of the data collectors’ training. Prior to deployment, the enumerators and supervisors received a 4-day training guided by a structured training manual. The training included an explanation of the sampling approach, basic principles of data collection, line-by-line discussion of the questionnaires, practicing the ODK system, training and standardization of anthropometric measurements, mock interviews, field practice, and a review of basic ethical practices of research involving human subjects.

### Indicators

There are multiple quality domains to consider, such as inputs (including service and equipment availability, training, etc.), service delivery processes (e.g., compliance with protocols and standards of care), and outcomes (e.g., health benefits, client satisfaction, etc.)[23]. For this study, we conducted an analysis of the readiness of health posts to manage wasting, focusing on four key attributes. This analysis was guided by the WHO’s Service Availability and Readiness Assessment, which encompasses facility infrastructure, staff guidelines, availability of equipment, and provision of medicine and nutrition commodities[24]. For process and intervention indicators, we used a quality of care checklist based on the national guidelines for the management of acute malnutrition classification algorithm and treatment protocols adapted to this study[25]. Our selection of domains and indicators for assessing the quality of care for wasting management was based on previous studies. We incorporated the domains that were commonly used in existing literature[7, 13].

**Table 1** provides an in-depth summary of the domains and indicators used to evaluate the quality of service and care for acute malnutrition. These domains were categorized into three primary sections: Service Readiness, Receipt of Complete Intervention, and Process Quality Care. The Receipt of Complete Intervention section consists of four indicators for SAM and three indicators for MAM, each designed to assess the comprehensiveness of interventions provided to children under 5 years of age diagnosed with malnutrition at health posts, with a distinct set of actions for each indicator.

**Table 1:** Summary of domains and indicators used to create indices of quality of service and care for acute malnutrition.

| Index | Domain | Number of indicators |  | Source of data |
| --- | --- | --- | --- | --- |
|  |  | SAM | MAM |  |
| Service readiness | Facility infrastructure | 2 | 2 | Facility |
|  | Staff guidelines | 7 | 8 | Facility |
|  | Equipment | 6 | 6 | Facility |
|  | Medicine and commodities | 7 | 3 | Facility |
| Receipt of complete intervention | Children under 5 who received appropriate treatment among children under age 5 who were diagnosed with malnutrition at the facility | 4 | 3 | Observation |
| Process quality care | Medical history | 5 | 5 | Observation |
|  | Physical examination | 6 | NA | Observation |
|  | Anthropometry measurement | 6 | 6 | Observation |
|  | Appetite Test | 7 | NA | Observation |
|  | Counselling | 10 | 8 | Observation |
|  | Recording | 4 | 5 | Observation |
|  | Follow up | 11 | 6 | Observation |

The process of quality of care in the management of acute malnutrition involves several critical categories that are essential for assessing the quality of service. We utilized five indicators to evaluate both Severe Acute Malnutrition (SAM) and Moderate Acute Malnutrition (MAM) for the medical history category. Similarly, we used five indicators for each SAM and MAM in the anthropometry measurement category. For physical examination, we used six indicators specifically for SAM. The appetite test category had seven indicators, all of which were specific to SAM. Counseling is a vital part of the care process, and we used ten indicators for SAM and eight for MAM. Recording is also an essential aspect of the process, and we used four indicators for SAM and five indicators for MAM. Finally, follow-up activities are a critical component of ongoing care and assessment, and we utilized 11 indicators for SAM and six for MAM.

This approach involves facilities reporting the presence of specific structures, commodities, and interviewers, confirming their availability and functionality on the survey day. Additionally, the observations included an assessment of whether appropriate interventions were provided in accordance with the national protocol for managing acute malnutrition. The indicators used to create the indices were binary. Each indicator was coded as either 1 if it was present (for structures/readiness) or had occurred (for processes and interventions), or 0 if it was not.

### Index creation

First, we developed composite indices based on the collected data, using a weighted additive approach. The weighted additive index was formed by summing the elements and considering the indicator count within each domain. This process involves adding the indicators in each domain, dividing the sum within each domain by the number of indicators in that specific domain, multiplying by 100, and finally dividing by the total number of domains within the index. Prior research has shown that using three different methods - simple additive, weighted additive, or principal component analysis - to compute quality indices typically produces results that are generally similar [26]. The weighted additive method has been recommended to be simple to calculate and interpret[27].

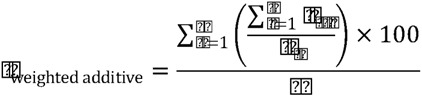

### Statistical analysis

We employed an ecological linking method to link health facilities and household survey data at the aggregate level. This approach has been demonstrated to be both feasible and valid for estimating effective coverage, while considering the quality of the services provided[28].

The effective coverage cascade for wasting management is calculated as follows:

*Quality-Adjusted Coverage = Contact Coverage× ReadinessX Receipt of Complete Intervention × Process Quality*

Where:

> *Coverage* was defined as the proportion of children who sought care at a health post among children under the age of five who were identified as malnourished in the household survey. *Readiness* was measured as the average readiness across health posts using the basic facility readiness index.
>
> *Receipt of Complete Intervention* is the proportion of children under 5 who received appropriate treatment among children under age 5 who were diagnosed with acute malnutrition at the facility. *Process Quality* is calculated as the average process quality across health posts using the basic process quality index for the management of wasting.

As described above, Effective Coverage (EC) is calculated by multiplying factors that signify both the readiness or effectiveness of services and the utilization of these services at the individual level The methodology employed for estimating effective coverage follows a cascading approach, as discussed in DHS methodological reports[29]. We combined data from four different files – household survey, service readiness, intervention, and process quality – into a single dataset using the “append” command in Stata.

We defined the new variable Y. In the household file, Y=contact coverage; in the readiness file, Y=mean service readiness score; in the intervention file, Y=mean receipt of complete intervention; in the process quality file, Y=mean process quality score. We extracted these four means as coefficients, constructed their products, and calculated the confidence interval for the product.

To obtain separate coefficients for Y in the four subfiles, we construct two dummy variables. In the household survey portion of the combined file, we defined x1=1, x2=0, x3=0, and x4=0. In the readiness portion of the file, x1=0, x2=1 x3=0 and x4=0. In the intervention portion of the file, x1=0, x2=0 x3=1 and x4=0. In the process-quality portion of the file, x1=0, x2=0 x3=0, and x4=1. We then applied a generalized linear model (GLM) without a constant term. Thus, the model is *glm Y x1 x2 x3 x4, family(binomial) link(logit) nocons*.

The resulting coefficients, namely b1 for coverage, b2 for readiness, b3 for receipt of complete intervention, and b4 for process quality, serve as pivotal indicators of these four key factors. The effective coverage estimates, denoted as EC, is then calculated as the product of these coefficients:

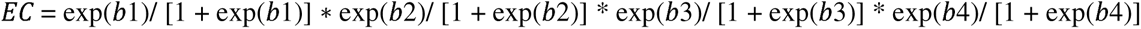

This formulation allows us to encapsulate the intricate interplay between coverage, readiness, receipt of a complete intervention, and process quality within the context of effective coverage estimation. The confidence intervals for cascaded effective coverage were obtained using the “nlcom” command in Stata. This command employs the delta method to automatically estimate the standard errors post-estimation[30]. A 95% confidence interval is calculated by adding or subtracting 1.96 times the standard error from the point estimate. Significant differences were determined using non-overlapping confidence intervals within at least two categories of disaggregation. Stata 17 was used for all analyses.

The data were disaggregated based on the following three variables:

### Wealth quintiles

In the household survey, each household was asked about ownership of a range of assets and housing materials. These responses were used to calculate a household wealth index. The wealth quintiles were calculated based on the distribution of the index across the surveyed population.

### Administrative Region

Each region is represented by two IMAM districts, as shown in **Supplementary Table 1.**

### Livelihood Region

Agrarian regions are represented by Amhara, Oromia, Sidama, and SNNP, whereas pastoral regions are represented by the Afar and Somali regions.

## Result

### Health facilities readiness

**Supplementary Table 2** provides insight into the preparedness of 72 health facilities in terms of the necessary structural components for managing SAM and MAM.

Regarding SAM management, approximately 40.3% of these health facilities had electricity, and 51.4% were equipped with an improved water source. Nearly three-fourths (73.6%) of the facilities possessed a quick translated reference guide for the outpatient therapeutic program (OTP). Additionally, 88.9% of these facilities had staff trained in diagnosing and managing acute malnutrition, both for SAM and MAM. While 59.7% of these institutions had lookup tables or charts, 90.3% maintained registration books for SAM treatment. Encouragingly, 95.8% had OTP Treatment and Follow-up Card. However, the availability of referral slips and guidelines or aids for Infant and Young Child Feeding counseling was limited, found in 11.1% and 47.2% of facilities, respectively.

The facilities were mostly equipped to manage SAM. Approximately 83.3% of them had scales for weighing children and infants, and 97.2% had MUAC tapes to measure the upper arm circumference of children under five years old. However, certain areas were lacking; only 37.5% had a dedicated weight scale, 66.7% had thermometers, and only 11.1% were equipped with WHO Growth charts. In addition, only 16.7% of the facilities had timers.

For medicines and commodities catering to SAM, 79.2% of the facilities were stocked with the RUTF. Other significant stocks included Vitamin A capsules in 77.8% of the facilities, Me-/albendazole cap/tablet (69.4 %), oral rehydration solution packets (68.1 %), and amoxicillin (55.6 %). Furthermore, 61.1% of the facilities were stocked with zinc sulfate tablets, whereas 58.3% stocked with IFA tablets. The readiness score for SAM management stood at 57.9%.

When examining the management of Moderate Acute Malnutrition (MAM), only approximately 59.7% of facilities had Ready-to-Use Supplementary Food (RUSF). Despite this, facilities were generally better equipped to manage MAM, with an overall readiness score of 76.4% (Supplementary Table 2).

### Receipt of complete intervention

**Supplementary Table 3** provides a detailed breakdown of the health facilities offering appropriate treatment and follow-ups for children under 5 diagnosed with acute malnutrition. This also included a score indicating the receipt of a complete intervention for each type of acute malnutrition.

In terms of SAM management, 69.5% of health providers correctly determined whether the child should be referred to SC or treated in the OTP, adhering to the action protocol. Routine medications, such as amoxicillin, were observed in 52.2% of patients. Furthermore, 65.2% of providers administered RUTF in proportion to the child’s weight. However, there appears to be room for improvement during follow-up visits; only 18.4% of health providers took appropriate actions or administered medications based on the child’s health condition during these visits. Considering all these factors, the overall score for the receipt of a complete intervention for SAM was 32.4%.

For Moderate Acute Malnutrition (MAM) management, providers provided routine medicines such as vitamin A and carried out deworming when required in 42.3% of the cases. RUSF was provided in line with national guidelines in 69.2% of the cases. However, during the follow-up visits, only 31.4% of the providers administered the necessary follow-up actions. The cumulative score for receipt of complete intervention in MAM cases was 41.8%.

### Process quality

**Supplementary Table 4** shows health facilities managing acute malnutrition in accordance with the recommended standards and presents their process quality scores.

### SAM Management

In the Medical History section, less than half of the providers (47.8%) asked about the history of diarrhea. Only about a quarter (26.1%) inquired about the child’s breastfeeding status and whether they had persistent cough. More than a third (34.8%) asked questions related to vomiting, and 39.1% checked whether the child had received all their routine vaccinations.

Under Physical Examination, 52.2% of the health providers checked for bilateral pitting edema. Temperature measurement was performed by 47.8%, while a mere 13.0% and 8.7% measured the respiratory rate and assessed dehydration, respectively. Only a few providers checked for skin lesions (4.3%) or checked the eyes and palms for signs of anemia (21.7%).

For anthropometric measurements, almost all providers (86.9 %) correctly measured the child’s MUAC and 82.6% accurately recorded and interpreted it. When considering the child’s weight, 69.6% of the providers both measured and recorded it accurately.

In the appetite test category, 26.1% of providers conducted the RUTF appetite test in a designated quiet area. Additionally, 21.7% of the providers ensured that hand washing occurred before feeding the RUTF and gently offered the child RUTF. Among the providers, 30.4% observed the child during feeding, 43.5% checked the amount consumed, and 56.5% determined the appetite test outcome.

The counseling practices varied. While 69.6% warned caregivers against sharing RUTF with others, only 13.1% specified not providing RUTF to infants aged 0-6 months. Approximately 43.5% of providers educated caregivers on the significance of breastfeeding for infants aged 6-23 months and the importance of feeding RUTF frequently. Almost half (47.8%) advised on the next visit, while 30.4% highlighted seeking immediate care if the child fell ill.

Most providers (86.9 %) completed OTP treatment and follow-up cards for recording purposes. Additionally, 95.6% of the providers correctly categorized patients on OTP admission. During Follow-ups, 87.7% checked MUAC and 81.6% verified their weight. Other checks, such as assessing dehydration or looking for skin lesions, were rarely performed. The total process quality care score for SAM was 53.2%.

### MAM Management

In relation to medical history, almost a third of the healthcare providers inquired about persistent diarrhea (30.8%) and vomiting (26.9%). Comparatively, fewer providers asked about extended cough or fever (7.7%), and approximately 19.2% inquired about their deworming history.

Anthropometric measurements and age determination were performed by 76.9% of the providers, whereas MUAC measurement, recording, and interpretation were impressively high, exceeding 88% in all these aspects. However, the correct measurement and recording of the child’s weight were notably lower.

During counseling sessions, 88.5% of caregivers were notified about their next visit. Additionally, 57.7% of providers advised against sharing RUSF with others, 7.7% advised not to provide it to infants less than six years of age and should be exclusively breastfed, and 34.6% emphasized its beneficial role in supplementing household diets and supporting breastfeeding.

For recording MAM cases, 96.1% registered the patient for MAM treatment and decided the appropriate TSFP admission category. In the Follow-up phase, weight was measured by 34.3%, and its gain or loss was recorded by 22.8%. Only a few checked for bilateral pitting edema (5.7%) or provided individual/group counseling (28.6%). The total process quality of care score for MAM was 37.4%.

### Components of Effective Coverage

**Supplementary Table 5** provides insights into the estimates of the various components of effective coverage measurement. These components encompass coverage, readiness, recall of a complete intervention, and process quality. The results are delineated by wealth quintile and region, for SAM and MAM.

The overall contact coverage for SAM was 40% (95% CI: 32, 48%). The readiness for SAM intervention was 58% (95% CI: 54, 62%). Furthermore, 32% (95% CI: 22, 42%) of the SAM cases received complete intervention. Additionally, the quality of the process was 53% (95% CI, 49–57%).

For the results across different wealth quintiles, it was observed that the contact coverage of SAM for the lowest-wealth group was 36% (95% CI: 14, 58%). The second quintile experienced slightly greater coverage at 69% (95% CI: 50%–88%). The middle, fourth, and highest quintiles had coverage rates of 34%, 35%, and 22%, respectively.

For the results across different regions, Afar SAM contact coverage at 36% (CI: 14, 58%), readiness was 57% (CI: 43, 71%), complete intervention receipt was 75% (CI: 46, 99%), and process quality was 65% (CI: 58, 73%). Similarly, in Amhara, the contact coverage was 30% (CI: 0.5, 34%), readiness was 71% (CI: 40, 79%), intervention receipt was 36% (CI: 7, 65%), and process quality was 69% (CI: 63, 75%). In Oromia, the contact coverage was 38% (CI: 8, 69%), readiness was 56% (CI: 46, 66%), intervention receipt was 6% (CI: 0.7, 18%), and process quality was 54% (CI: 51, 57%). Sidama showed a high contact coverage of 76% (CI: 58, 94%) and readiness of 63% (CI: 55, 71%), but lower intervention receipt and process quality of 25% (CI: 7, 43%) and 44% (CI: 36–51%), respectively. For SNNP, coverage was 33% (CI: 2 65%), readiness was 53% (CI: 46, 61%), intervention receipt was 11% (CI: 0.7, 2%), and process quality was 26% (CI: 19, 33%). Finally, in the Somali region, the coverage was 26% (CI: 13, 39%), readiness was 46% (CI: 39%, 53%), intervention receipt was 42% (CI: 9, 74%), and process quality was 61% (CI: 51, 71%).

The MAM intervention had an overall contact coverage of 37% (CI: 31, 42%), readiness score of 76% (CI: 72, 81%), and completion rate of 42% (CI: 31, 53%). In terms of wealth quintiles, the lowest quintile had the highest contact coverage at 55% (CI: 41, 69%), whereas the second, middle, fourth, and highest quintiles had coverage rates of 34% (CI: 19, 49%), 38% (CI: 25, 51%), 28% (CI: 19, 38%), and 32% (CI: 17, 48%), respectively.

Across the regions, Afar had 44% MAM contact coverage (CI: 29, 59%), Amhara had 36% (CI: 21, 52%), Sidama had 58% (CI: 45, 72%), SNNP had 52% (CI: 30, 74%), and Somali had 13% (CI: 5, 21%). Regarding readiness, Afar readiness score was 77% (CI: 62, 91%), Amhara 94% (CI: 89, 98%), Oromia 67% (CI: 54, 80%), and Somali 62% (CI: 50, 74%). 65% (CI: 43, 88%) of children in Afar received complete intervention, while 29% (CI: 0.6, 58%) did in Sidama, 16% (CI: 1, 32%) in SNNP, and 75% (CI: 50, 99%) in Amhara. The process quality score was 24% (CI: 8, 40%) in SNNP, 48% (CI: 34, 62%) in Oromia, 28% (CI: 13, 43%) in Sidama, 46% (CI: 30, 63%) in Afar, and 40% (CI: 23, 56%) in Amhara.

**Figure 1** depicts a visual representation of treatment coverage, along with various other metrics, including readiness, completion of the entire intervention, and the quality of the process. In the case of SAM, the contact coverage was below 50% in most regions, except for Sidama, whereas all regions, except Somali, had readiness scores above 50%. For MAM, the treatment coverage was below 50% in all regions except for Sidama and SNNP, but the readiness scores were consistently above 50%. The complete intervention rate for SAM was below 50% in all regions except Afar, whereas the MAM complete intervention rate was above 50% only in Afar and Amhara. The process quality scores for SAM were above 50% in all regions, except SNNP and Sidama, whereas for MAM, all regions had process quality scores below 50%.

**Figure 1:**
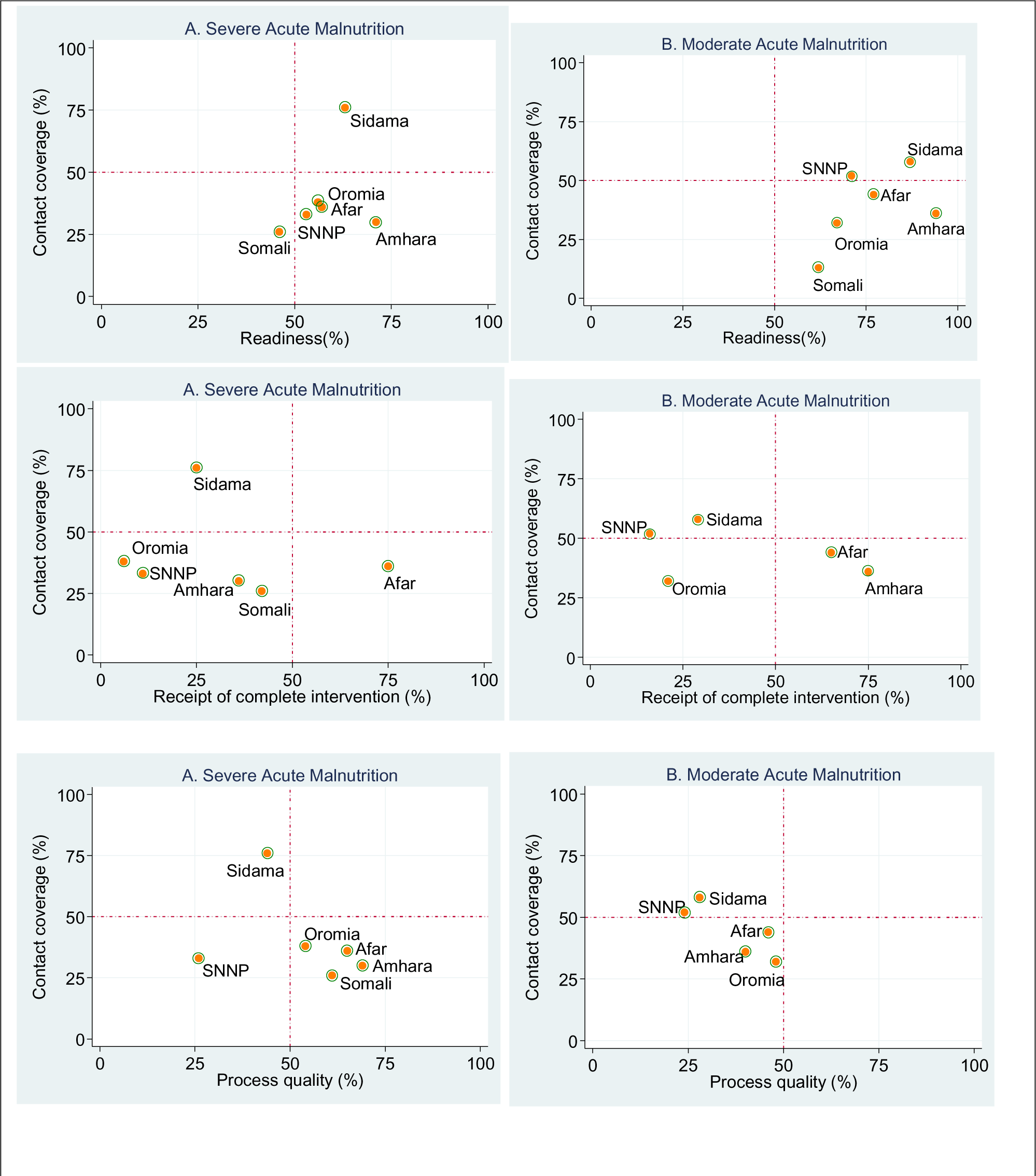
Regional readiness, receipt of complete intervention, and process quality score of facilities to provide nutrition interventions versus coverage in Ethiopia in 2023.

When the metrics are aggregated at the livelihood region level, contact coverage for pastoralist regions was below 50% for SAM, while agrarian regions coverage was above 50%. Furthermore, both regions exhibited readiness scores exceeding 50%. For MAM, both regions have contact coverage below 50%, but readiness scores are higher than 50%. The complete intervention for SAM is below 50% in agrarian regions, and the same is true for MAM in these regions. Additionally, process quality scores for SAM were above 50% in pastoral regions, whereas both regions had scores below 50% for MAM (Supplementary Figure 1).

### Effective Coverage Cascade

**Figure 2** shows the effective coverage measurements for both the SAM and MAM. Contact coverage was reported to be 40% (95% CI: 32, 48%). The input-adjusted coverage was 23% (95% CI: 16, 30%), intervention-adjusted coverage was 7% (95% CI: 4, 11%), and quality-adjusted coverage was 4% (95% CI: 2, 6%). Likewise, the contact coverage was 37% (95% CI: 31%, 42%). The input-adjusted coverage, intervention-adjusted coverage, and quality-adjusted coverage were 28% (95% CI: 22, 34%), 12% (95% CI: 8, 16%), and % (95% CI: 3, 6%), respectively.

**Figure 2:**
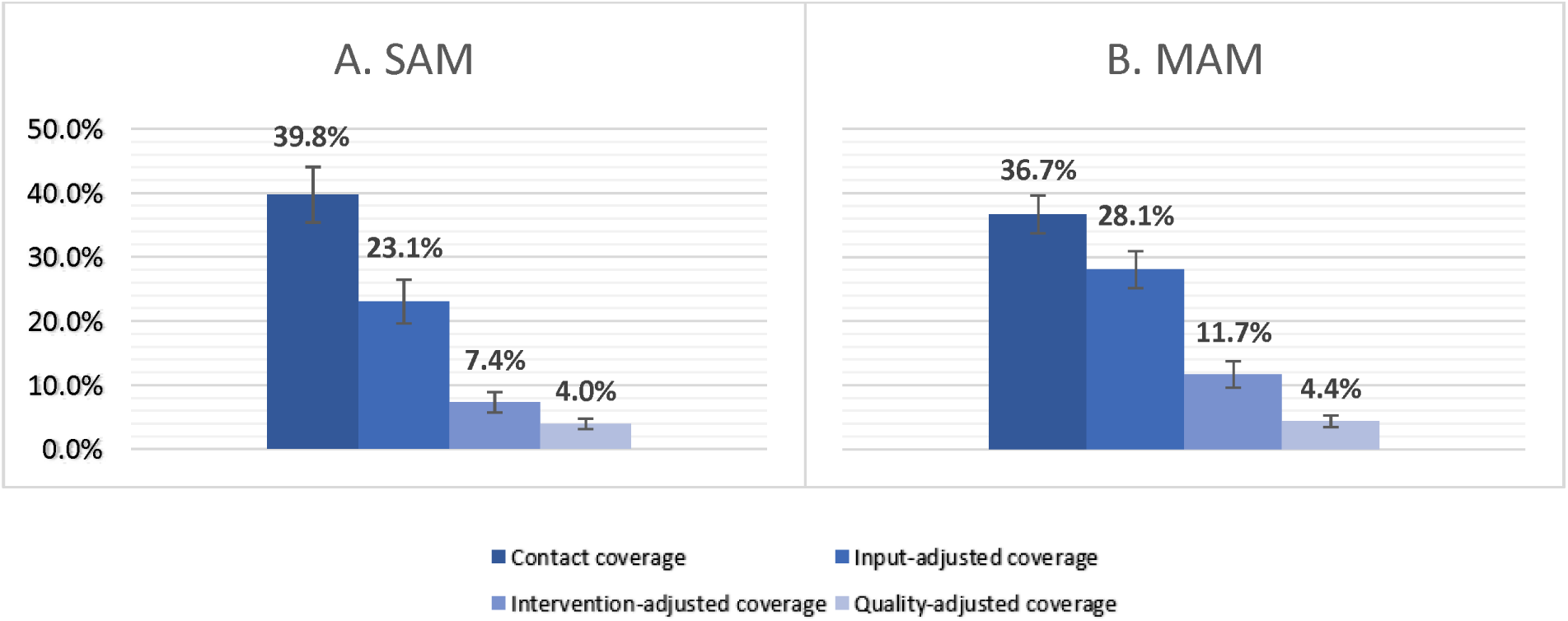
SAM and MAM Effective Coverage Cascade, Ethiopia, 2023.

Figure 3 displays the effective coverage measurements for both SAM and MAM stratified by wealth quintiles. For SMA, the contact coverage was highest in the second quintile at 69% (95% CI: 50, 88%), followed by the lowest quintile at 42% (95% CI: 9, 74%), and gradually decreased to 22% (95% CI: 5, 39%) for the highest quintile. The middle and fourth quintiles had contact coverages of 34% (95% CI: 16, 53%) and 35% (95% CI: 18, 52%), respectively. The input-adjusted, intervention-adjusted, and quality-adjusted coverage across these quintiles exhibited a similar pattern of variability. In the case of MAM, the lowest quintile had the highest contact coverage rate of 55% (95% CI: 41, 69%). In contrast, the second, middle, fourth, and highest quintiles had contact coverage rates of 34% (95% CI: 19, 49%), 38% (CI: 25, 51%), 28% (95% CI: 19, 38%), and 32% (95% CI:17, 48%), respectively. The coverage values across the quintiles were variable and followed a similar pattern when adjusted for inputs, interventions, and quality.

**Figure 3:**
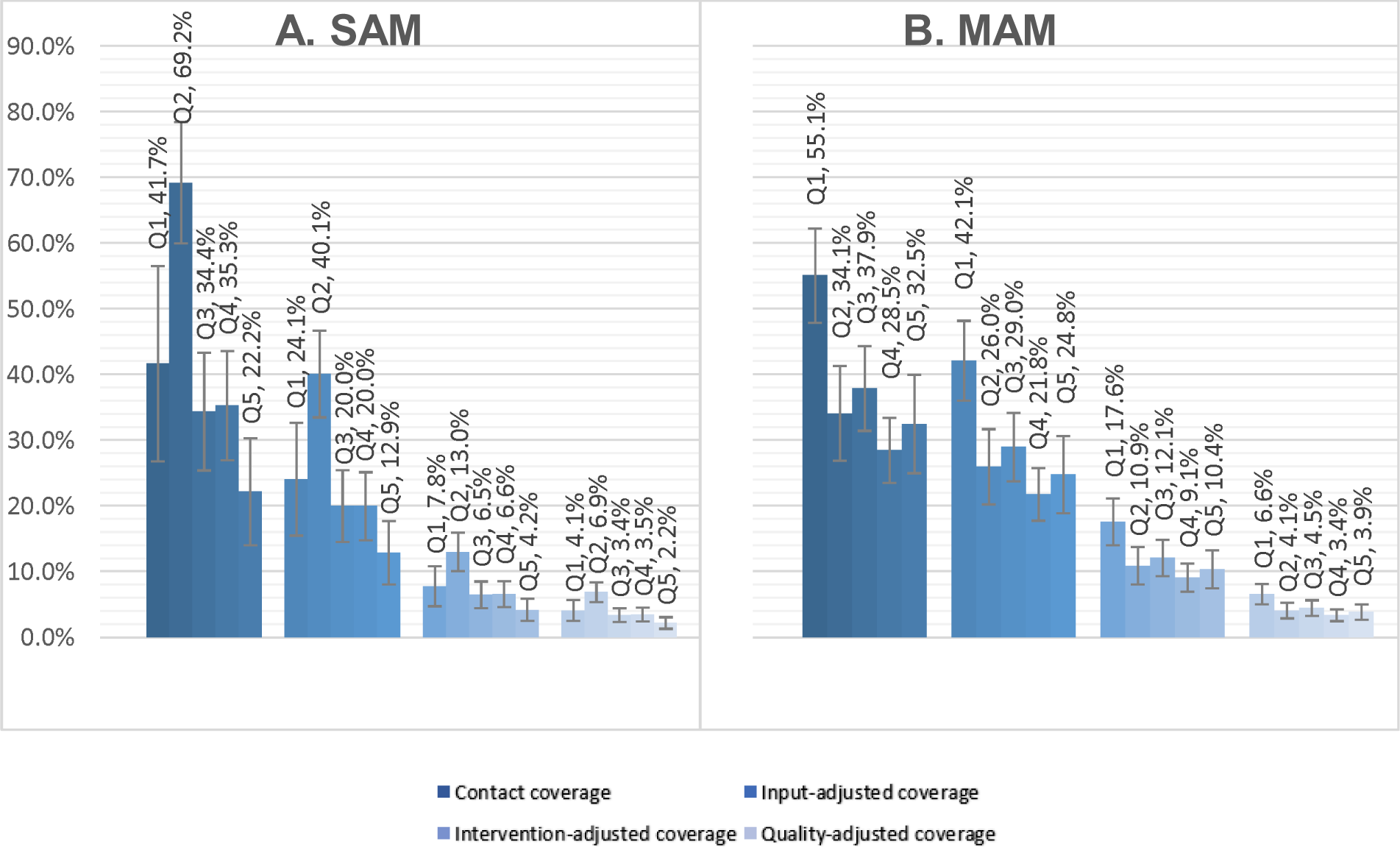
SAM and MAM Effective Coverage by Wealth Index, Ethiopia, 2023.

Figure 4 depicts the effective coverage measurements for SAM and MAM by region. With respect to SAM, the Sidama Region shows significantly higher coverage compared to other, especially when compared with the Somali Region at 76% (95% CI: 58, 94%). Once more, Sidama Region shows the highest input-adjusted coverage, significantly higher than Somali Region at 48% (95% CI: 25, 71%). In the case of intervention-adjusted coverage, no significant difference found among regions. In terms of quality-adjusted coverage, Afar Region shows the highest coverage at 10% (95% CI: 2, 18%), significantly higher than SNNP region. Similarly, in comparison to other regions, Sidama Region demonstrates a higher contact coverage rate of 58% (95% CI: 45, 72%) for MAM, with a particularly significant difference when compared with the Somali Region. Sidama Region also shows the highest input-adjusted coverage, significantly higher than Somali and Oromia Regions at 51% (95% CI: 35, 66%). In the case of intervention and quality adjusted coverages, no significant difference was found among regions. When the metrics are aggregated at the livelihood region level, there were no significant differences among agrarian and pastoralist regions (as shown in **Supplementary Figure 2**).

**Figure 4:**
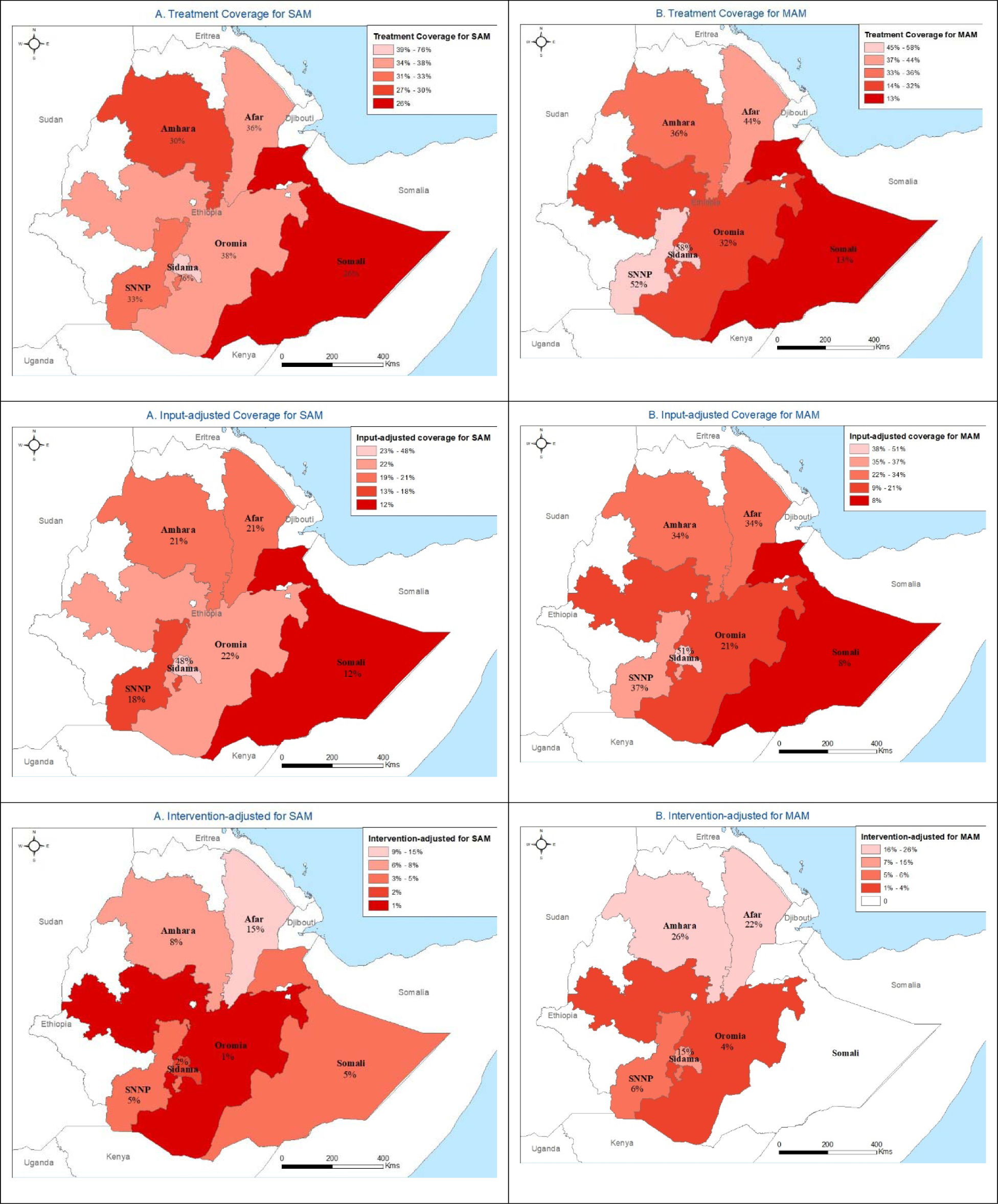
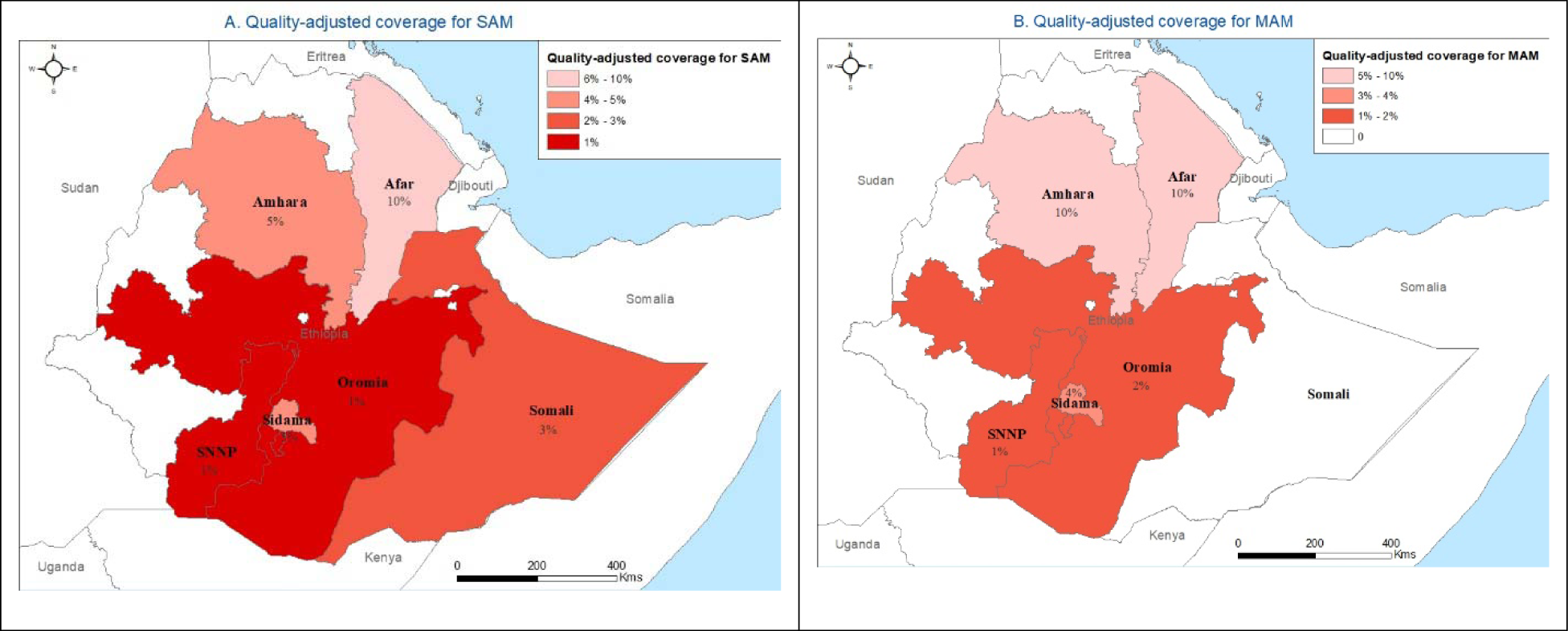
Contact coverage, input-adjusted coverage, intervention-adjusted coverage, and quality-adjusted coverage for severe and moderate acute malnutrition services by region, Ethiopia 2023.

## Discussion

Child malnutrition remains a pressing public health concern, with Acute Malnutrition being the predominant contributor to child morbidity and mortality in various low-income and middle-income countries[1]. Effective management and intervention in healthcare facilities play a crucial role in addressing acute malnutrition. Our findings show differentiated readiness among health facilities: the readiness score for SAM management stood at 57.9%, while for MAM, it was higher at 76.4%. This indicates greater preparedness for managing MAM than SAM does. Regarding interventions, only 32.4% of SAM cases received complete intervention compared to a slightly higher 41.8% in MAM cases. Process quality of care also varied, with the total process quality care score for SAM cases being 53.2%, whereas MAM cases had a score of 37.4%.

In terms of SAM coverage, the contact coverage was 40%. Upon further examination of the quality-adjusted measures, more concerning figures emerge input-adjusted coverage was only 23%, intervention-adjusted coverage was as low as 7%, and quality-adjusted coverage was only 4%. The MAM intervention displayed slightly different dynamics, with a total contact coverage of 37%, input-adjusted coverage of 28%, intervention-adjusted coverage of 12%, and quality-adjusted coverage of 4%.

Results from cluster surveys conducted in five African countries[31] and a comprehensive analysis from a study focused on the coverage of community-based management of severe acute malnutrition programmes across twenty-one countries [32] echo a worrisome trend: the treatment coverage for SAM is far from satisfactory. The former revealed a coverage of less than 20%, while the latter indicated that only 38.3% of community-based SAM programs achieved the minimum standards of the SPHERE project. Rural programs reported a meager coverage rate of 34.6 %. Our data from Ethiopia align with this narrative, with a contact coverage of 40% for SAM.

In the Ethiopian context, this situation resonates with broader trends. A study in Ethiopia highlighted that while the country has made remarkable progress in expanding its primary healthcare infrastructure, the quality of its services lags. The country’s health system struggles with challenges ranging from shortages of essential supplies and trained personnel to irregularities in care provision[33]. Thus, the findings from the present study align with the prevailing narrative, reiterating the imperative need for bolstered efforts towards enhancing the quality of care while ensuring its accessibility.

Our findings are in contrast to those presented in a recent evaluation of nutrition-specific interventions in Ethiopia[21]. A key difference lies in the metrics used to measure coverage and quality. The study gauged coverage at the facility level, determining delivery based on the recording of both admissions and discharges of children under five years of age. Quality in their assessment was shaped by evaluations of nutrition supply management, health workers’ knowledge, interactions between health workers and beneficiaries, and beneficiaries’ satisfaction with the delivered nutrition service. While these metrics offer insights into delivery services, they may not delve deeply into the complexities of care quality as in our approach. We adopted a more nuanced set of metrics that provided a detailed look into the intricacies of SAM and MAM care delivery and quality. The disparities between our findings emphasize the importance of understanding and interpreting data in the context of the methodologies and metrics employed.

The readiness of health facilities for wasting management presents a mixed picture, with some areas showing potential and others highlighting the significant gaps essential for comprehensive malnutrition care. The presence of quick reference guides in 73.6% of the facilities and trained staff in 88.9% of them underscores the concerted efforts put forth to bolster the knowledge base and training in these facilities. Compared to a study in Mozambique, where 58.2% of health professionals received nutrition-related training[34], our study showed a more pronounced emphasis on staff training. This can potentially be attributed to the recent waves of training in the new national guidelines for managing acute malnutrition in Ethiopia[21].

However, although knowledge and training are indispensable, they are only one side of the coin. The tangible infrastructure that facilitates the translation of this knowledge into an effective action remains a challenge. For instance, the lack of essential amenities such as electricity in 59.7% of the health facilities was concerning. Further complicating the scenario is the insufficient access to improved water sources in nearly half of the health facilities. The importance of clean water cannot be overemphasized, especially when discussing nutritional interventions. Clean water is vital for procedures such as the appetite test, which is a cornerstone of SAM management. The findings of studies in Ethiopia that detailed gaps in structural readiness align with this observation, signaling that infrastructural readiness lags[35, 36].

In our study, a significant number of facilities had high availability rates of RUTF (79.2%) and RUSF (59.7%), suggesting an effective supply chain for supporting nutritional interventions. Compared with the findings of Esete et al., in which 75% of the surveyed health facilities had essential supplies such as the RUTF[35], the results align, further emphasizing the overarching efficiency of the supply chain. However, this benchmark standard of care seems somewhat disparate when considering our observed complete intervention score for SAM of just 32.4%. This is further accentuated by the fact that, even when RUTF and RUSF were accessible, only 65% and 69% of patients, respectively, received them according to the standard guidelines. The depth of these gaps becomes even more pronounced when considering the continuity of care: only 18% of SAM cases benefited from vital follow-up actions, such as home visits, a number that slightly improved to 31% for MAM cases. This shortcoming underscores that the provision of therapeutic foods, while imperative, is merely a component of a broader continuum of care that should seamlessly integrate follow-ups, monitoring, counseling, and other elements of holistic care [37]. This suggests potential bottlenecks in the actual delivery of care, or other barriers that prevent optimal usage. Conforming to the World Health Organization guidelines for severe acute malnutrition care can potentially reduce case fatality rates by 41% [38].

This narrative, a combination of commendable achievements in resource availability yet unfulfilled potential in comprehensive care delivery, is consistent with broader concerns in the healthcare domain in many developing contexts [39], including Ethiopia[33]. The gaps observed between having resources and optimally utilizing them emphasize the need for a more integrative approach to SAM and MAM management, ensuring that every child not only has access to medicines, but also benefits from a comprehensive care plan that holistically addresses their health needs. Esete et al. illuminates a tangible manifestation of this issue, showing that critical contact points like immunization units present missed opportunities. A mere 16.4% of children were given anthropometric assessments, and only 16.2% of mothers with children under six months of age received counseling about exclusive breastfeeding at these points[35].

Efforts to strengthen wasting management are evident, with quick reference guides present in 73.6% of facilities and 88.9% of trained staff. However, the practical implementation of this training has exhibited inconsistencies. Gaps existed in areas such as appetite tests (26.1%) and physical examinations (ranging from 4.3% to 52.2%). Suboptimal sensitivity in detecting malnutrition among health extension workers in Ethiopia was also noted, starting at 34% and modestly improving to 48% by endline[40]. The results emphasize that merely implementing guidelines, offering training, and providing essential equipment does not necessarily guarantee that care is delivered according to established standards [41–45]. It is worth noting that assessments of nutritional services in other settings have also highlighted poor adherence to guidelines, leading to substandard health outcomes[42, 45, 46].

Our study revealed a significant disparity between contact-based and quality-adjusted coverage. While the initial contact coverage for SAM and MAM was 40% and 37%, respectively, it plummeted to a mere 4% after quality adjustment. This noteworthy decrease highlights the critical importance of delivering high-quality care, rather than focusing solely on reaching patients (Supplementary Table 6). Our results align with previous research that has shown that effective coverage is often much lower than contact coverage for antenatal care (ANC), family planning, and sick childcare in several low- and middle-income countries (LMICs)[29, 39, 47, 48].

Moreover, our study revealed disparities in coverage metrics across wealth quintiles, emphasizing the socioeconomic barriers to healthcare access and quality. Both SAM and MAM exhibit substantial disparities in coverage across wealth quintiles, with the highest wealth quintiles having lower coverage than those in the lower quintiles. This contradicts evidence showing an increase in care-seeking for sick children as wealth increases [49]. Consequently, it becomes evident that ensuring access to healthcare alone is insufficient. Identifying areas of substandard care represents a vital step towards advancing healthcare services and bridging the gaps that hinder effective healthcare provision[45, 50–53].

Regional breakdown revealed substantial variations, suggesting geographical disparities in the management of wasting. The data indicate that agrarian regions have higher coverage across all measures than pastoralist regions; however, quality-adjusted coverage remains low in both types of regions. The provision of free maternal and child interventions in governmental health facilities in Ethiopia may contribute to inequities, primarily because of limited access to healthcare services because of remote locations, low utilization of health services, and/or poor quality of healthcare services[10].

Addressing disparities in access to and quality of care is of paramount importance, especially in disadvantaged populations. Poor quality care can result in several negative outcomes, such as unnecessary health-related suffering, persistent symptoms, loss of function, and lack of trust and confidence in health systems. Additionally, poor-quality health systems can lead to a waste of resources and catastrophic expenditures[54].

Our study has several strengths. First, household surveys and facility data were collected in the same year, enabling a comparison between the preparedness of facilities and the services provided for acute malnutrition at the same point in time. Second, our selection of items for readiness and process quality measures was guided by the WHO’s Service Availability and Readiness Assessment and the National Guideline for the Management of Acute Malnutrition, which was adapted from the WHO guidelines. We also assigned equal weights to each item within a measure, aligning with recommendations from previous research on the measurement of quality of care [55]. Finally, we adopted a more nuanced set of metrics that provided a detailed look into the intricacies of SAM and MAM care delivery and quality.

This study had several limitations. Firstly, data on the wealth quintiles were only available from household surveys. Although this methodology is influenced by data availability, it does not completely capture the variations in readiness or process quality experienced by individuals with specific characteristics when visiting a particular health facility.[48]. Second, our analysis was constrained by the selective inclusion of health posts in IMAM districts. This limitation affects the generalizability of our findings, as the characteristics and quality of care in health centers and hospitals may differ significantly from those observed in health posts. Thus, nuances in patient management of severe acute malnutrition, specifically those services available at the health center and hospital levels, were not observed in our study. Finally, despite the comprehensiveness of the health facility survey, it falls short in capturing data essential for holistically calculating the care cascade from the need for services to health benefits. Notably, this study lacks the necessary information for computing user adherence-adjusted coverage or outcome-adjusted coverage[7, 12].

In conclusion, the Ethiopian health system’s efforts to address acute child malnutrition show a mix of commendable strides and critical gaps. Despite training and resource availability, practical application in managing malnutrition has revealed inconsistencies. The challenge extends beyond merely bringing malnourished children to health facilities to ensure that they receive holistic, high-quality care. While significant progress is evident, a more integrated approach, synergizing resources with consistent, quality-driven interventions, is crucial to effectively combat wasting in Ethiopia.

## Acknowledgements

The authors wish to thank the study data collectors and district and health facility staff.

## Author contribution

AM, SH conceived the study. AM analyzed the data and wrote the first draft with inputs from SH, SG. All authors have read and approved the final manuscript.

## Ethical consideration

The baseline and health facilities surveys were designed and carried out with the utmost respect for the rights of study participants, upholding principles of research ethics such as justice, beneficence, and non-maleficence. This study was conducted in compliance with both national and international ethical guidelines, with approval from the Institutional Review Board (IRB) of the College of Health Sciences, AAU. Necessary administrative clearances were obtained from the Ministry of Health (MoH) and relevant regional health bureaus. Data were collected after obtaining informed consent from both parents and caregivers of children aged 6-59 months and pregnant women (PLW). To ensure beneficence, children and PLW who met the program admission criteria but had not been admitted were linked with the program.

## Data availability statement

The dataset for this study is available from the corresponding author on reasonable request.

## Map disclaimer

The depiction of boundaries on this map does not imply the expression of any opinion whatsoever on the part of BMJ (or any member of its group) concerning the legal status of any country, territory, jurisdiction, or area or of its authorities. This map is provided without any warranty of any kind, either express or implied.

---

**Supplementary table 1:** Districts included in the study.

| Regions | IMAM + IS | IMAM – IS | Non-IMAM districts |
| --- | --- | --- | --- |
| Afar | Mille | Chifra | Adaar |
| Amhara | Dalanta | Meket | Dawunt |
| Oromia | Girawa | Bedeno | Deder |
| Somali | Adadle | Mustahil | Gode |
| Sidama | Aleta Chuko | Shebedino | Dale |
| SNNP | Zala | Uba Dhebresahay | Kucha |

SNNP=Southern National and Nationality People, IMAM=Integrated Management of Acute Malnutrition, IS=Implementation Science.

**Supplementary table 2:** Percentage of health facilities with structural input items and their readiness score.

| Indicator | % facilities with items available (95% CI) |
| --- | --- |
| **Severe Acute Malnutrition (SAM) Management (n=72 facilities)** |  |
| Facility infrastructure (2) |  |
| Electricity | 40.3 (28.8, 51.9) |
| Improved water source | 51.4(39.6, 63.2) |
| Staff and guidelines (7) |  |
| Quick translated reference guide for OTP and TSFP | 73.6(63.2, 84.0) |
| Staff trained in diagnosis and management of acute malnutrition (SAM and MAM) | 88.9(81.4, 96.3) |
| Look up tables/charts available | 59.7(48.1, 71.3) |
| Registration book for SAM treatment | 90.3(83.3, 97.3) |
| Outpatient Therapeutic Programme (OTP) Treatment and Follow-up Card | 95.8(91.1, 100) |
| Referral slip | 11.1(3.7, 18.5) |
| Infant and Young Child Feeding counselling guideline /job-aids/quick reference. | 47.2(35.4, 59.0) |
| Equipment (6) |  |
| Child and infant scale | 83.3(74.5, 92.2) |
| Weight scale | 37.5(26.0, 48.9) |
| Thermometer | 66.7(55.5, 77.8) |
| MUAC tapes for U5 children | 97.2(93.3, 100) |
| Growth charts (WHO) | 11.1(3.7, 18.5) |
| Timer | 16.7(7.8, 25.5) |
| Medicine and commodities |  |
| RUTF | 79.2(69.5, 88.8) |
| Oral rehydration solution packet | 68.1(57.0, 79.1) |
| Amoxicillin (dispersible tablet 250 or 500 mg OR syrup/suspension) | 55.6(43.8, 67.3) |
| Vitamin A capsules | 77.8(67.9, 87.6) |
| Me-/albendazole cap/tab | 69.4(58.5, 80.3) |
| Zinc sulphate tablets, dispersible tablets, or syrup | 61.1(49.6, 72.6) |
| IFA tablets | 58.3(46.7, 67.0) |
| **Readiness score for SAM management** | **57.9(54.1, 61.7)** |
| **Moderate Acute Malnutrition (MAM) Management (n=72 facilities)** |  |
| Facility infrastructure (2) |  |
| Electricity | 40.3 (28.8, 51.9) |
| Improved water source | 51.4(39.6, 63.2) |
| Staff and guidelines (8) |  |
| Quick translated reference guide for OTP and TSFP | 73.6(63.2, 84.0) |
| Staff trained in diagnosis and management of acute malnutrition (SAM and MAM) | 88.9(81.4, 96.3) |
| Look up tables/charts available | 59.7(48.1, 71.3) |
| Registration book for MAM treatment (U5) | 86.1(77.9, 94.3) |
| MAM Treatment and Follow-up Card | 88.9(81.4, 96.3) |
| TSFP Identification Card | 77.8(67.9, 87.6) |
| Referral slip | 11.1(3.7, 18.5) |
| Infant and Young Child Feeding counselling guideline /job-aids/quick reference. | 47.2(35.4, 59.0) |
| Equipment (6) |  |
| Child and infant scale | 83.3(74.5, 92.2) |
| Weight scale | 37.5(26.0, 48.9) |
| Thermometer | 66.7(55.5, 77.8) |
| MUAC tapes for U5 children | 97.2(93.3, 100) |
| Growth charts (WHO) | 11.1(3.7, 18.5) |
| Timer | 16.7(7.8, 25.5) |
| Medicine and commodities (3) |  |
| RUSF | 59.7(48.1, 71.3) |
| Vitamin A capsules | 77.8(67.9, 87.6) |
| Me-/albendazole cap/tab | 69.4(58.5, 80.3) |
| **Readiness score for MAM management** | **76.4(71.6, 81.1)** |

**Supplementary Table 3:**
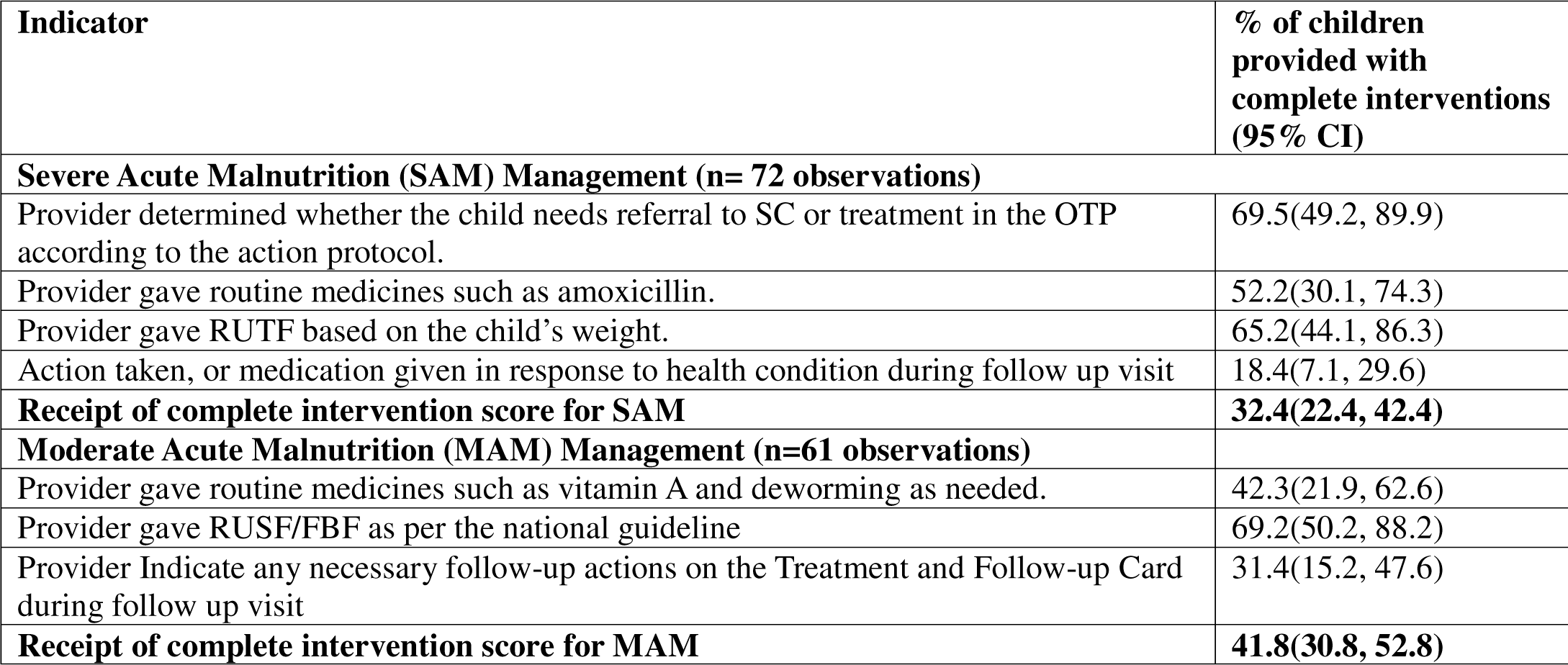
Health facilities providing appropriate treatment and follow-up among U5 children diagnosed with acute malnutrition and their receipt of complete intervention score.

**Supplementary Table 4:** Health facilities providing acute malnutrition services according to recommended standards and process quality scores.

| Indicator | % of facilities provided process quality of care (95% CI) |
| --- | --- |
| **Severe Acute Malnutrition (SAM) Management** |  |
| Medical history (5) |  |
| Provider asked If the child has diarrhea >14 days. | 47.8(25.7, 69.9) |
| Provider asked if the child for breastfeeding status | 26.1(6.7, 45.5) |
| Provider asked if the child had a cough > 14 days. | 26.1(6.7, 45.5) |
| Provider asked If the child had vomiting | 34.8(10.2, 55.8) |
| Provider asked If the child completed routine Immunization | 39.1(17.5, 60.7) |
| Physical examination (6) |  |
| Provider checked for presence of bilateral pitting edema | 52.2(30.1, 74.3) |
| Provider measured temperature | 47.8(25.7, 69.9) |
| Provider measured respiratory rate | 13.0(0.18, 27.1) |
| Provider assessed for dehydration | 8.7(0.37, 21.1) |
| Provider examined for skin lesions | 4.3(0.04, 13.4) |
| Provider checked for eye/palms for signs of anemia | 21.7(3.5, 39.9) |
| Anthropometry measurement (6) |  |
| Provider determined age | 1 |
| Provider correctly measured the child's weight | 69.6(49.2, 89.9) |
| Provider correctly recorded the child's weight | 69.6(49.2, 89.9) |
| Provider correctly measured MUAC | 86.9(72.1, 99.8) |
| Provider correctly record MUAC | 82.6(65.8, 99.3) |
| Provider correctly interpreted MUAC | 82.6(65.8, 99.3) |
| Appetite Test (7) |  |
| Provider conducted the RUTF appetite test in a quiet, separate area | 26.1(6.7, 45.5) |
| Provider explained to the caregiver the purpose and procedure of the appetite test. | 39.1(17.5, 60.7) |
| Provider asked the mother/caregiver to wash hands with soap before feeding the RUTF | 21.7(3.5, 39.9) |
| Asked the mother/caregiver to gently offer the child RUTF | 21.7(3.5, 39.9) |
| Provider observed the child eating the RUTF. | 30.4(10.1, 50.7) |
| Provider checked the amount of RUTF consumed by the child. | 43.5(21.6, 65.4) |
| Provider determined whether he/she passes or fails the appetite test. | 56.5(34.6, 78.4) |
| Counselling (10) |  |
| Provider informed the caregiver not to give RUTF to infants 0-6 months and they should be exclusively breastfed. | 13.1(1.8, 27.9) |
| Provider informed the caregiver not Share RUTF to other children or family members in the households. | 69.6(49.2, 89.9) |
| Provider informed the caregiver if the child is 6-23 months to continue to breastfeed on demand. Mother's milk is best for infants and young children. | 43.5(21.6, 65.4) |
| Provider informed the caregiver to feed the child small amounts of RUTF until the allocated daily ration is finished, and before eating other food apart from breast milk. | 34.8(13.7, 55.8) |
| The provider informed the caregiver to encourage the child to eat as often as possible (every 3 hours during the day). | 43.5(21.6, 65.4) |
| Provider informed the caregiver If the child is breastfeeding, to offer breast milk on demand and before feeding RUTF. | 21.7(3.5, 39.9) |
| Provider informed the caregiver to provide plenty of safe drinking water for the child while eating RUTF. | 34.8(13.7, 55.8) |
| Provider informed the caregiver not to mix RUTF with liquids | 34.8(13.7, 55.8) |
| Provider advised the caregiver or patient when to return for the next visit. | 47.8(25.7, 69.9) |
| Provider advised the caregiver to seek medical care at the nearest health facility immediately if child becomes ill while at home or refuses to eat RUTF. | 30.4(10.1, 50.8) |
| **Recording (4)** |  |
| Provider completed the OTP Treatment and Follow-up Card. | 86.9(72.1, 99.9) |
| Provider registered the patient in the registration book for SAM treatment. | 1 |
| Provider decided which OTP admission entry category to assign to the patient. | 95.6(86.6, 99.9) |
| Provider assigned a registration number if the patient is a new admission. | 1 |
| **Follow up (11)** |  |
| Provider asked if the child has suffered any illness since the last visit | 44.9(30.5, 59.3) |
| Provider checked MUAC | 87.7(78.2, 97.3) |
| Provider checked weight | 81.6(70.4, 92.9) |
| Provider checked for degree of bilateral pitting oedema | 30.6(17.2, 44.0) |
| Provider recorded child's weight and MUAC | 75.5(63.0, 88.0) |
| Provider measured temperature (axillary temp) | 34.7(20.9, 48.5) |
| Provider measured respiratory rate | 1 |
| Provider assessed dehydration | 4.1(0.02, 9.8) |
| Provider examined for skin lesions | 2.0(0.02, 6.1) |
| Provider examined eye/palm for signs of anemia | 1 |
| Provider conducted appetite test | 32.6(19.0, 46.3) |
| **Total process quality care score for SAM** | **53.2(48.9, 57.4)** |
| **Moderate Acute Malnutrition (MAM) Management** |  |
| **Medical history (5)** |  |
| Provider asked If the child has diarrhea >14 days. | 30.8(11.7, 49.8) |
| Provider asked if the child had a cough > 14 days. | 7.7(0.03, 18.7) |
| Provider asked If the child had vomiting | 26.9(8.6, 45.2) |
| Provider asked if the child had fever | 7.7(0.03, 18.7) |
| Provider asked history of deworming | 19.2(3.0, 35.5) |
| **Anthropometry measurement (6)** |  |
| Provider determined age | 76.9(59.5, 94.3) |
| Provider correctly measured the child's weight | 19.2(3.0, 35.5) |
| Provider correctly recorded the child's weight | 30.8(11.7, 49.8) |
| Provider correctly measured MUAC | 88.5(75.3, 99.9) |
| Provider correctly record MUAC | 96.1(88.2, 99.9) |
| Provider correctly interpreted MUAC | 96.1(88.2, 99.9) |
| **Counselling (8)** |  |
| The provider informed the caregiver not to give RUSF to infants 0-6 months and they should be exclusively breastfed. | 7.7(0.03, 18.7) |
| Provider informed the caregiver not Share RUSF to other children or family members in the households. | 57.7(37.3, 78.0) |
| Provider informed the caregiver RUSF should be consumed in addition to, and not as a substitute for, the household diet and breastfeeding. | 34.6(15.0, 54.2) |
| Provider informed the caregiver If the child is breastfeeding, to offer breast milk on demand and before feeding RUSF. | 26.9(8.6, 45.2) |
| Provider informed the caregiver to provide plenty of safe drinking water for the child while eating RUSF. | 26.9(8.6, 45.2) |
| Provider informed the caregiver RUSF is ready to be used, therefore should not be mixed with other foods. | 30.1(11.7, 49.8) |
| The provider informed the caregiver once opened, store the RUSF sachet in a clean, cool, and dry place. | 26.9(8.6, 45.2) |
| Provider advised the caregiver or patient when to return for the next visit. | 88.5(75.3, 99.9) |
| Recording (5) |  |
| Provider completed the TSFP Treatment and Follow-up Card. | 1 |
| Provider registered the patient in the registration book for MAM treatment. | 96.1(88.2, 99.9) |
| Provider decided which TSFP admission entry category to assign to the patient. | 96.1(88.2, 99.9) |
| Provider assigned a registration number if the patient is a new admission. | 1 |
| Provider issue TSFP identification card to the patient | 88.5(75.3, 99.9) |
| Follow up (6) |  |
| Provider measured weight | 34.3(17.7, 50.8) |
| Provider recode wight gain/loss | 22.8(8.2, 37.5) |
| Provider checked for degree of bilateral pitting oedema | 5.7(0.02, 13.8) |
| Provider inform the caregiver or patient on the progress of treatment. | 25.7(10.5, 40.9) |
| Provider conduct individual and/or group counselling on health, WASH, and IYCF | 28.6(12.8, 44.3) |
| **Total process quality of care score for MAM** | **37.4(30.8, 44.0)** |

\*1=100%

**Supplementary Table 5:**
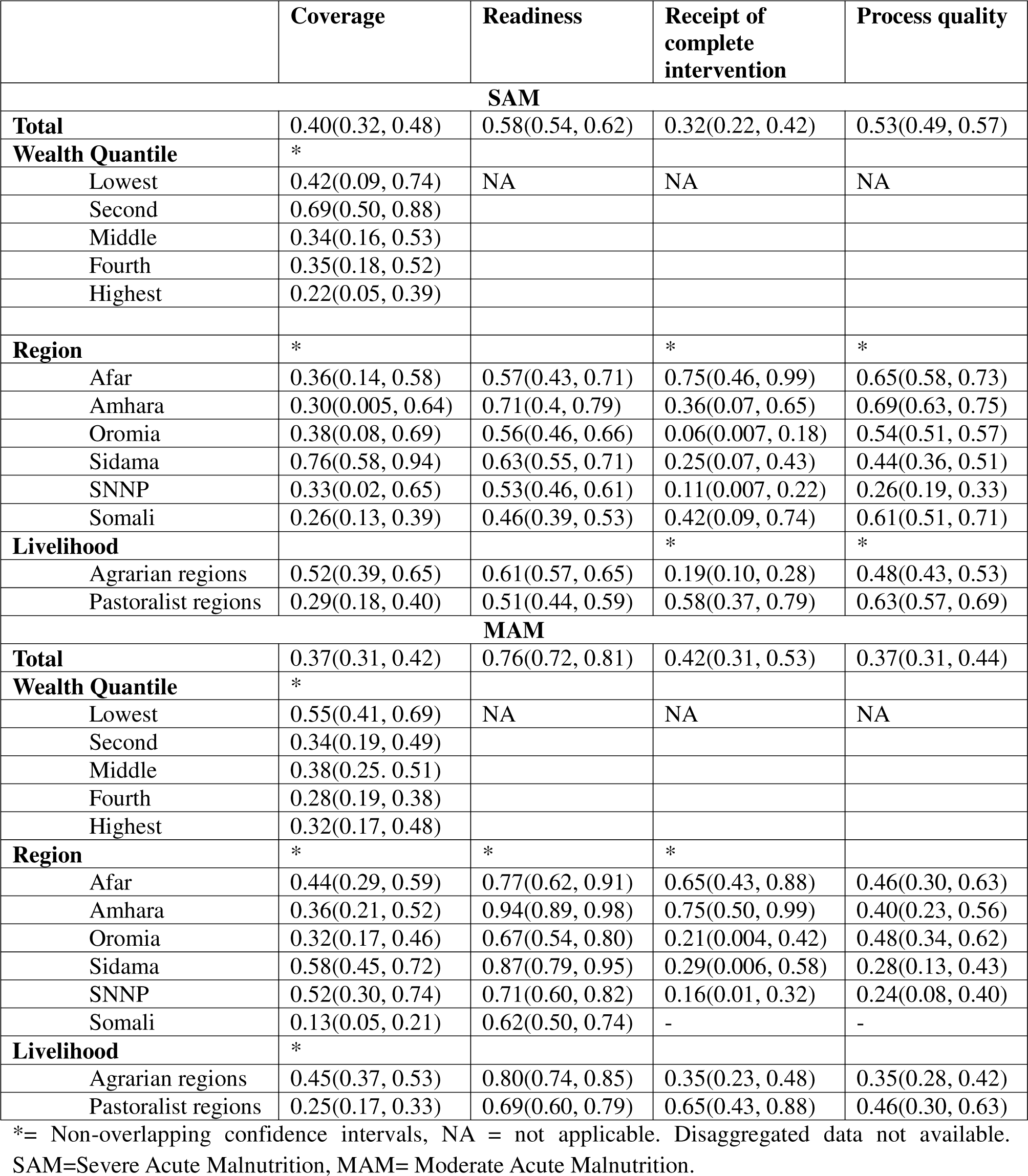
Estimates of each component of effective coverage measurement by wealth quintile and region. Proportions with 95% confidence intervals.

|  | Coverage | Readiness | Receipt of complete intervention | Process quality |
| --- | --- | --- | --- | --- |
| **SAM** |  |  |  |  |
| **Total** | 0.40(0.32, 0.48) | 0.58(0.54, 0.62) | 0.32(0.22, 0.42) | 0.53(0.49, 0.57) |
| **Wealth Quintile** | * |  |  |  |
| Lowest | 0.42(0.09, 0.74) | NA | NA | NA |
| Second | 0.69(0.50, 0.88) |  |  |  |
| Middle | 0.34(0.16, 0.53) |  |  |  |
| Fourth | 0.35(0.18, 0.52) |  |  |  |
| Highest | 0.22(0.05, 0.39) |  |  |  |
| **Region** | * |  | * | * |
| Afar | 0.36(0.14, 0.58) | 0.57(0.43, 0.71) | 0.75(0.46, 0.99) | 0.65(0.58, 0.73) |
| Amhara | 0.30(0.005, 0.64) | 0.71(0.4, 0.79) | 0.36(0.07, 0.65) | 0.69(0.63, 0.75) |
| Oromia | 0.38(0.08, 0.69) | 0.56(0.46, 0.66) | 0.06(0.007, 0.18) | 0.54(0.51, 0.57) |
| Sidama | 0.76(0.58, 0.94) | 0.63(0.55, 0.71) | 0.25(0.07, 0.43) | 0.44(0.36, 0.51) |
| SNNP | 0.33(0.02, 0.65) | 0.53(0.46, 0.61) | 0.11(0.007, 0.22) | 0.26(0.19, 0.33) |
| Somali | 0.26(0.13, 0.39) | 0.46(0.39, 0.53) | 0.42(0.09, 0.74) | 0.61(0.51, 0.71) |
| **Livelihood** |  |  | * | * |
| Agrarian regions | 0.52(0.39, 0.65) | 0.61(0.57, 0.65) | 0.19(0.10, 0.28) | 0.48(0.43, 0.53) |
| Pastoralist regions | 0.29(0.18, 0.40) | 0.51(0.44, 0.59) | 0.58(0.37, 0.79) | 0.63(0.57, 0.69) |
| **MAM** |  |  |  |  |
| **Total** | 0.37(0.31, 0.42) | 0.76(0.72, 0.81) | 0.42(0.31, 0.53) | 0.37(0.31, 0.44) |
| **Wealth Quintile** | * |  |  |  |
| Lowest | 0.55(0.41, 0.69) | NA | NA | NA |
| Second | 0.34(0.19, 0.49) |  |  |  |
| Middle | 0.38(0.25, 0.51) |  |  |  |
| Fourth | 0.28(0.19, 0.38) |  |  |  |
| Highest | 0.32(0.17, 0.48) |  |  |  |
| **Region** | * | * | * |  |
| Afar | 0.44(0.29, 0.59) | 0.77(0.62, 0.91) | 0.65(0.43, 0.88) | 0.46(0.30, 0.63) |
| Amhara | 0.36(0.21, 0.52) | 0.94(0.89, 0.98) | 0.75(0.50, 0.99) | 0.40(0.23, 0.56) |
| Oromia | 0.32(0.17, 0.46) | 0.67(0.54, 0.80) | 0.21(0.004, 0.42) | 0.48(0.34, 0.62) |
| Sidama | 0.58(0.45, 0.72) | 0.87(0.79, 0.95) | 0.29(0.006, 0.58) | 0.28(0.13, 0.43) |
| SNNP | 0.52(0.30, 0.74) | 0.71(0.60, 0.82) | 0.16(0.01, 0.32) | 0.24(0.08, 0.40) |
| Somali | 0.13(0.05, 0.21) | 0.62(0.50, 0.74) | - | - |
| **Livelihood** | * |  |  |  |
| Agrarian regions | 0.45(0.37, 0.53) | 0.80(0.74, 0.85) | 0.35(0.23, 0.48) | 0.35(0.28, 0.42) |
| Pastoralist regions | 0.25(0.17, 0.33) | 0.69(0.60, 0.79) | 0.65(0.43, 0.88) | 0.46(0.30, 0.63) |

\*= Non-overlapping confidence intervals, NA = not applicable. Disaggregated data not available. SAM=Severe Acute Malnutrition, MAM= Moderate Acute Malnutrition.

**Supplementary Table 6:** Estimates of each component of the effective coverage measurement by wealth quintile and region. Proportions with 95% confidence intervals.

|  | Contact coverage | Input-adjusted coverage | Intervention-adjusted coverage | Quality-adjusted coverage |
| --- | --- | --- | --- | --- |
| **SAM** |  |  |  |  |
| **Total** | 0.40(0.32, 0.48) | 0.23(0.16, 0.30) | 0.07(0.04, 0.11) | 0.04(0.02, 0.06) |
| **Wealth Quintile** | * | * |  |  |
| Lowest | 0.42(0.09, 0.74) | 0.24(0.07, 0.41) | 0.08(0.02, 0.14) | 0.04(0.01, 0.07) |
| Second | 0.69(0.50, 0.88) | 0.40(0.27, 0.53) | 0.13(0.07, 0.19) | 0.07(0.04, 0.10) |
| Middle | 0.34(0.16, 0.53) | 0.20(0.09, 0.31) | 0.06(0.02, 0.10) | 0.03(0.01, 0.05) |
| Fourth | 0.35(0.18, 0.52) | 0.20(0.10, 0.30) | 0.07(0.03, 0.10) | 0.04(0.02, 0.06) |
| Highest | 0.22(0.05, 0.39) | 0.13(0.03, 0.22) | 0.04(0.009, 0.07) | 0.02(0.005, 0.04) |
| **Region** | * | * |  | * |
| Afar | 0.36(0.14, 0.58) | 0.21(0.05, 0.36) | 0.15(0.04, 0.27) | 0.10(0.02, 0.18) |
| Amhara | 0.30(0.005, 0.64) | 0.21(0.003, 0.43) | 0.08(0.0002, 0.17) | 0.05(0.0009, 0.11) |
| Oromia | 0.38(0.08, 0.69) | 0.22(0.03, 0.40) | 0.01(0.0001, 0.04) | 0.01(0.0005, 0.02) |
| Sidama | 0.76(0.58, 0.94) | 0.48(0.25, 0.71) | 0.12(0.006, 0.24) | 0.05(0.005, 0.10) |
| SNNP | 0.33(0.02, 0.65) | 0.18(0.007, 0.35) | 0.02(0.001, 0.06) | 0.005(0.0003, 0.01) |
| Somali | 0.26(0.13, 0.39) | 0.12(0.03, 0.21) | 0.05(0.003, 0.10) | 0.03(0.003, 0.06) |
| **Livelihood** |  |  |  |  |
| Agrarian regions | 0.52(0.39, 0.65) | 0.31(0.21, 0.42) | 0.06(0.02, 0.10) | 0.03(0.01, 0.05) |
| Pastoralist regions | 0.29(0.18, 0.40) | 0.15(0.07, 0.23) | 0.09(0.04, 0.14) | 0.06(0.02, 0.09) |
| **MAM** |  |  |  |  |
| **Total** | 0.37(0.31, 0.42) | 0.28(0.22, 0.34) | 0.12(0.08, 0.16) | 0.04(0.03, 0.06) |
| **Wealth Quintile** | * |  |  |  |
| Lowest | 0.55(0.41, 0.69) | 0.42(0.03, 0.54) | 0.18(0.11, 0.25) | 0.07(0.04, 0.10) |
| Second | 0.34(0.19, 0.49) | 0.26(0.15, 0.37) | 0.11(0.05, 0.16) | 0.04(0.02, 0.06) |
| Middle | 0.38(0.25, 0.51) | 0.29(0.19, 0.39) | 0.12(0.07, 0.18) | 0.05(0.02, 0.07) |
| Fourth | 0.28(0.19, 0.38) | 0.22(0.14, 0.30) | 0.09(0.05, 0.13) | 0.03(0.02, 0.05) |
| Highest | 0.32(0.17, 0.48) | 0.25(0.13, 0.36) | 0.10(0.05, 0.16) | 0.04(0.02, 0.06) |
| **Region** | * | * |  |  |
| Afar | 0.44(0.29, 0.59) | 0.34(0.19, 0.50) | 0.22(0.10, 0.35) | 0.10(0.03, 0.18) |
| Amhara | 0.36(0.21, 0.52) | 0.34(0.20, 0.49) | 0.26(0.12, 0.39) | 0.10(0.02, 0.19) |
| Oromia | 0.32(0.17, 0.46) | 0.21(0.09, 0.34) | 0.04(0.0009, 0.10) | 0.02(0.0002, 0.04) |
| Sidama | 0.58(0.45, 0.72) | 0.51(0.35, 0.66) | 0.15(0.01, 0.28) | 0.04(0.0004, 0.09) |
| SNNP | 0.52(0.30, 0.74) | 0.37(0.17, 0.57) | 0.06(0.0002, 0.14) | 0.01(0.0006, 0.03) |
| Somali | 0.13(0.05, 0.21) | 0.08(0.02, 0.14) | - | - |
| **Livelihood** | * | * |  |  |
| Agrarian regions | 0.45(0.37, 0.53) | 0.36(0.28, 0.44) | 0.13(0.07, 0.18) | 0.04(0.02, 0.07) |
| Pastoralist regions | 0.25(0.17, 0.33) | 0.17(0.10, 0.25) | 0.11(0.05, 0.17) | 0.05(0.01, 0.09) |

\*= Non-overlapping confidence intervals, SAM=Severe Acute Malnutrition, MAM= Moderate Acute Malnutrition,

**Supplementary Figure 1:**
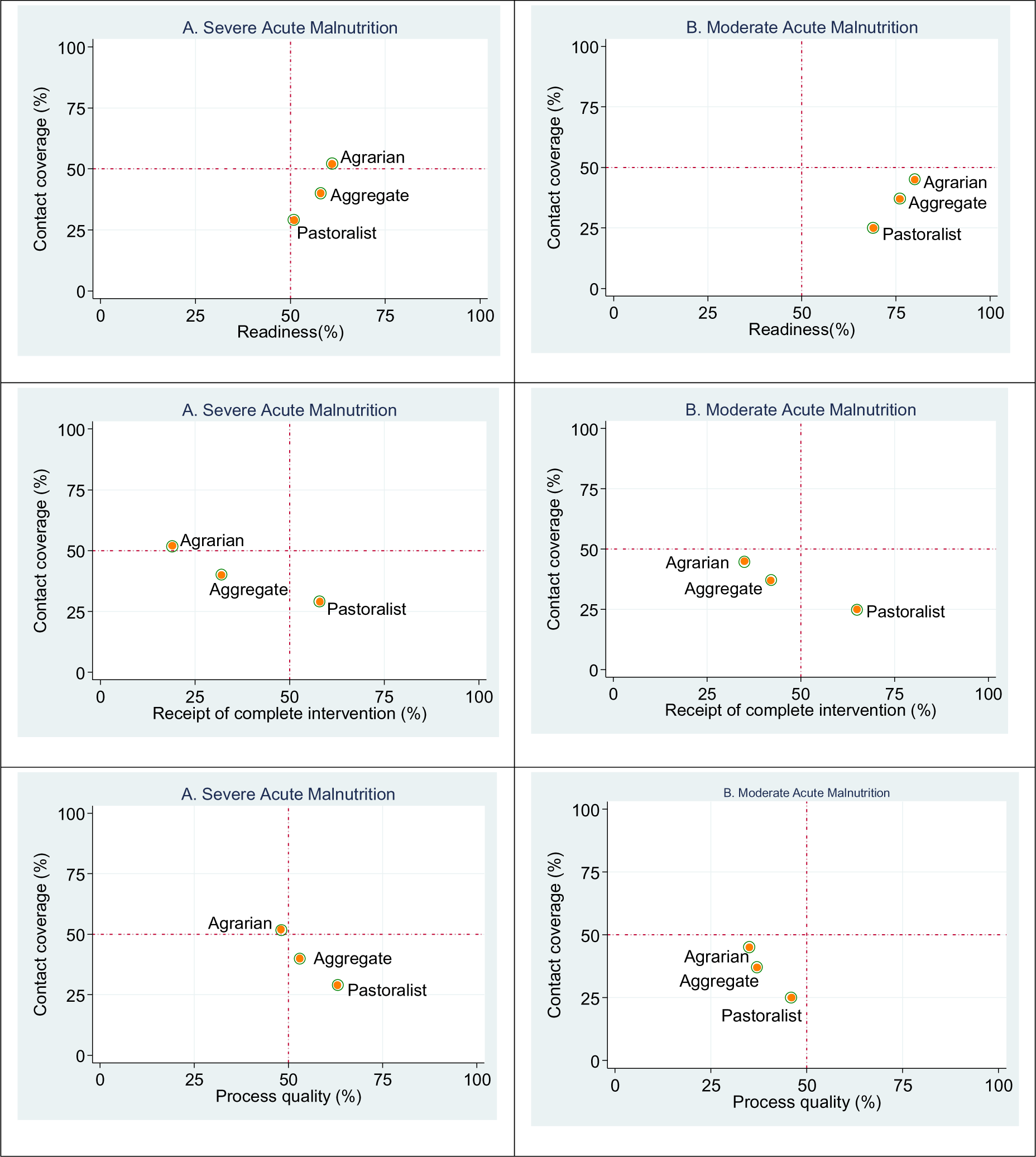
Livelihood regional readiness, receipt of complete intervention, and process quality score of facilities to provide nutrition interventions versus coverage, Ethiopia, 2023.

**Figure 2:**
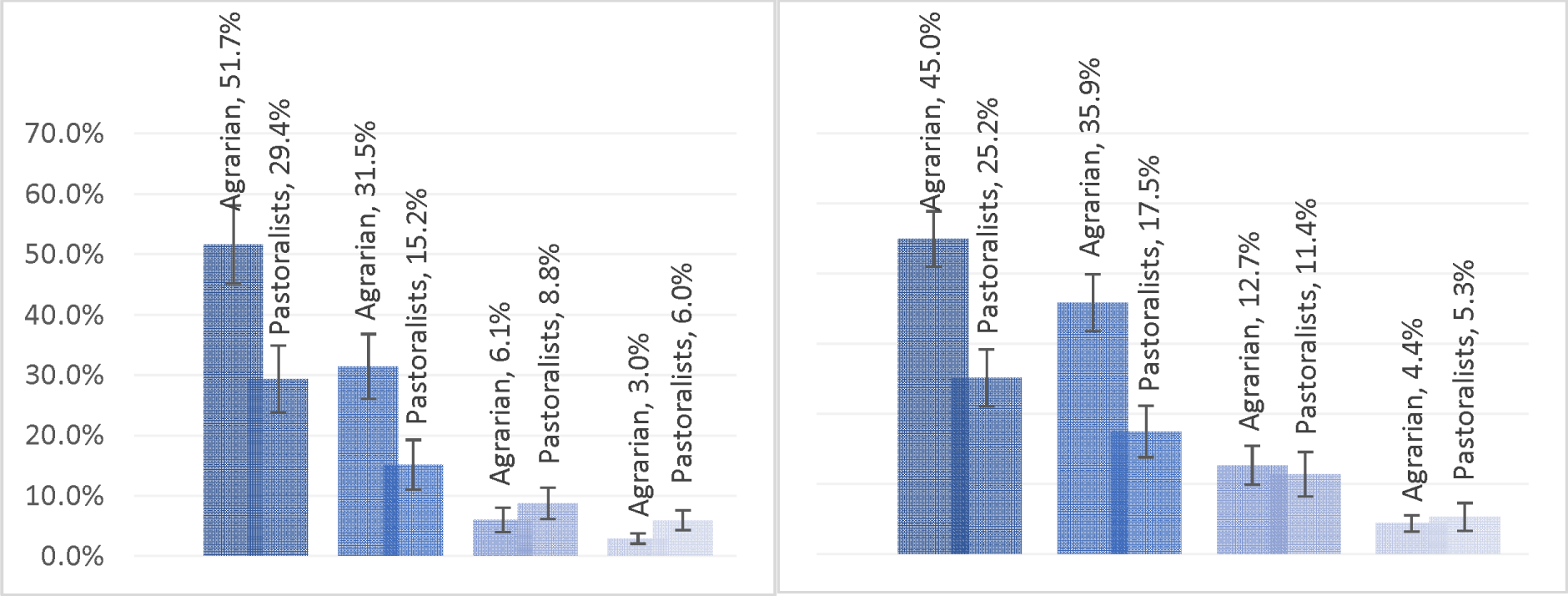
Effective coverage of SAM and MAM by livelihood regions, Ethiopia 2023.

